# Multimodality Quantitative Assessment of Left Atrial Remodeling Characteristics Using Echocardiography and LGE-CMR in Isolated Degenerative Mitral Regurgitation: Association With Atrial Fibrillation

**DOI:** 10.64898/2026.07.31.26359443

**Authors:** Xinmu Li, Yanyan Song, Yiran Hu, Lingyu Mi, Leyi Zhu, Fengwen Zhang, Fang Fang, Minjie Lu, Xiangbin Pan

## Abstract

**Background:** Although atrial fibrillation (AF) frequently coexists with degenerative mitral regurgitation (DMR), imaging assessment of left atrial (LA) fibrosis in this population remains limited. We integrated echocardiographic and cardiac magnetic resonance (CMR) late gadolinium enhancement (LGE) parameters to characterize LA remodeling features in isolated DMR and assess its association with AF.

**Methods:** In this retrospective study, consecutive patients with moderate-to-severe or severe isolated DMR who underwent echocardiography and CMR, including cine and LGE sequences, between February 2025 and January 2026 were included. Patients with other left-sided valvular disease of mild or greater severity or cardiomyopathy were excluded to isolate a population of “pure” DMR patients, and LALGE percentage was calculated as LALGE area divided by left atrial wall area. Imaging parameters were compared according to AF status and evaluated using logistic regression and receiver operating characteristic (ROC) analysis. Multimodality models were constructed to assess the combined discriminative value of echocardiographic and CMR parameters.

**Results:** Among 46 patients with DM (71.7% male, 62.0 years of age), 17 (37.0%) had AF. Compared with patients without AF, AF patients were older (*P*=0.005) and had lower LA reservoir strain [LASr; adjusted odds ratio (OR) 0.86; 95% confidence interval (CI) 0.76-0.94; *P*<0.001], higher left atrioventricular coupling index (LACI; adjusted OR per 10-percentage-point increase, 1.48; 95% CI, 1.18-2.23; *P*<0.001), larger LALGE area (adjusted OR 1.20; 95% CI, 1.07-1.41; *P*<0.001), higher LALGE percentage (adjusted OR 1.19; 95% CI, 1.02-1.42; *P*=0.025). Among single parameters, LACI showed the highest area under the curve (AUC 0.85, 95% CI 0.72-0.99; cutoff 58.59%), whereas LALGE area showed an AUC of 0.78 (95% CI 0.61-0.94; cutoff 13.73 cm²). Among all models, LASr combined with LALGE percentage achieved the highest AUC (0.91, 95% CI 0.80-1.00), with sensitivity 76.5%, specificity 96.6%, positive predictive value 92.9%, and negative predictive value 87.5%. In serum biomarker analysis, patients with AF had significantly higher levels of C-terminal telopeptide of type I collagen (CITP) than those without AF (*P*=0.009). Follow-up data were available for 45 patients (97.8%), with a median follow-up duration of 9.2 months (IQR, 7.7-12.2 months).

**Conclusions:** In patients with DMR, AF was associated with more advanced LA structural remodeling, impaired LA function, and higher CMR LGE burden. Echocardiographic LA functional parameters, when combined with LALGE measures, may provide complementary information for the discrimination of AF. Given the exploratory nature of this study, the findings require external validation.

## Background

Degenerative mitral regurgitation (DMR), caused by primary abnormalities of the mitral valve apparatus such as leaflet prolapse or flail, affects at least 24 million people worldwide.^1^ Mechanistically, DMR differs from functional mitral regurgitation (FMR), which occurs secondary to left ventricular remodeling and/or mitral annular dilatation in the absence of primary leaflet pathology.^2–4^ Despite their distinct mechanisms, left atrial (LA) remodeling is common across the spectrum of MR and may contribute to atrial fibrillation (AF) development and maintenance. AF is among the most common arrhythmias in clinical practice and represents a major source of cardiovascular morbidity and mortality, affecting approximately 10.5 million adults in the United States.^5–7^ Although AF frequently coexists with DMR in clinical practice, its diagnosis, prognostic implications, and management in this population remain relatively underemphasized in contemporary reviews and guidelines.^8,9^ In patients with DMR, AF may represent not only a rhythm disorder but also a marker of advanced atrial remodeling, reflecting the cumulative burden of structural, functional, and fibrotic changes within the atrial myocardium.^10,11^

Despite advances in imaging and surgical strategy, clinical monitoring and management of DMR patients with AF remain challenging.^12^ Conventional clinical assessment mainly relies on symptom status, LA size, pulmonary pressure, and left ventricular function; however, these parameters may not fully capture the complexity of atrial myocardial dysfunction and fibrosis.^13,14^ A detailed characterization of LA remodeling in DMR and its association with AF may help refine risk stratification, inform the timing of mitral valve intervention, and improve clinical decision-making.^15^

Echocardiography and cardiac magnetic resonance (CMR) provide complementary information on LA remodeling. Echocardiography allows quantitative assessment of LA size and function, such as the left atrioventricular coupling index (LACI) and left atrial reservoir strain (LASr), which reflect LA structural-functional remodeling and have demonstrated prognostic value in cardiovascular disease.^13,16^ Three-dimensional high-resolution late gadolinium enhancement CMR allows noninvasive visualization and quantification of LA fibrosis, providing additional information on atrial tissue remodeling that may not be captured by conventional echocardiographic measurements.^14,17^ Nevertheless, the combined value of echocardiographic functional indices and CMR-derived LA late gadolinium enhancement in isolated DMR remains incompletely defined.

Therefore, this study integrated quantitative echocardiographic and CMR parameters to characterize structural, functional, and fibrotic LA remodeling in patients with isolated DMR and to evaluate their associations with AF. These findings may provide further insight into the interaction between valvular heart disease and atrial remodeling, and may help identify imaging markers associated with advanced atrial substrate in DMR.

## Methods

### Study Population

In this retrospective observational study, consecutive patients with moderate-to-severe or severe isolated DMR evaluated between February 2025 and January 2026 were included if they were aged ≥18 years and underwent both transthoracic echocardiography and CMR, including cine and late gadolinium enhancement (LGE) sequences suitable for quantitative assessment of LALGE. Exclusion criteria were any form of cardiomyopathy, previous surgical or catheter ablation for AF before enrollment, concomitant mild or greater stenosis or regurgitation of other left-sided valves, and inadequate CMR image quality precluding reliable LALGE analysis.

Patients were classified into the AF group and non-AF group according to their baseline AF status. AF diagnosis was determined based on 12-lead electrocardiography, 24-hour Holter monitoring, inpatient medical records, or outpatient records. Patients with a documented history of previous or current AF were assigned to the AF group. Clinical and demographic data, including age, sex, comorbidities, cardiac rhythm, and mitral valve intervention strategies were collected at enrollment. The patients were followed up from hospital discharge. Follow-up outcomes included all-cause mortality, heart failure hospitalization, stroke, postoperative or new-onset atrial fibrillation, AF-related procedures, and recurrent mitral regurgitation. Recurrent MR was defined as MR ≥2+ after initially successful surgical repair and as MR ≥3+ after initially successful transcatheter edge-to-edge repair, whereas new or worsening pathological prosthetic mitral regurgitation was evaluated separately after mitral valve replacement.^18–20^ Follow-up data were obtained from electronic medical records, outpatient evaluations, and telephone interviews.

The study was conducted in accordance with the Declaration of Helsinki and approved by the Ethics Committee of Fuwai Hospital, Chinese Academy of Medical Sciences (No. 2025-2621). The written informed consent was obtained.

### Echocardiography

Echocardiographic examinations were performed using a Vivid E95 ultrasound system (GE HealthCare, Norway) and images were analyzed offline using TomTec-Arena workstation (version 2.30; Philips Healthcare, Germany). Echocardiographic assessment included LA and left ventricular structure and function, mitral valve morphology, and the severity of valvular regurgitation. LA measurements included transverse and longitudinal diameters, maximum LA volume (LAVmax), minimum LA volume (LAVmin), LA ejection fraction (LAEF), LA sphericity index, LA reservoir strain (LASr, **Figure 1**), LA conduit strain (LAScd), left atrioventricular coupling index (LACI), LA-to-left ventricular volume ratio (LA:LV), and LA stiffness index (LASI). LAEF was calculated as:

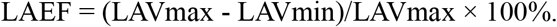

**Figure 1.**
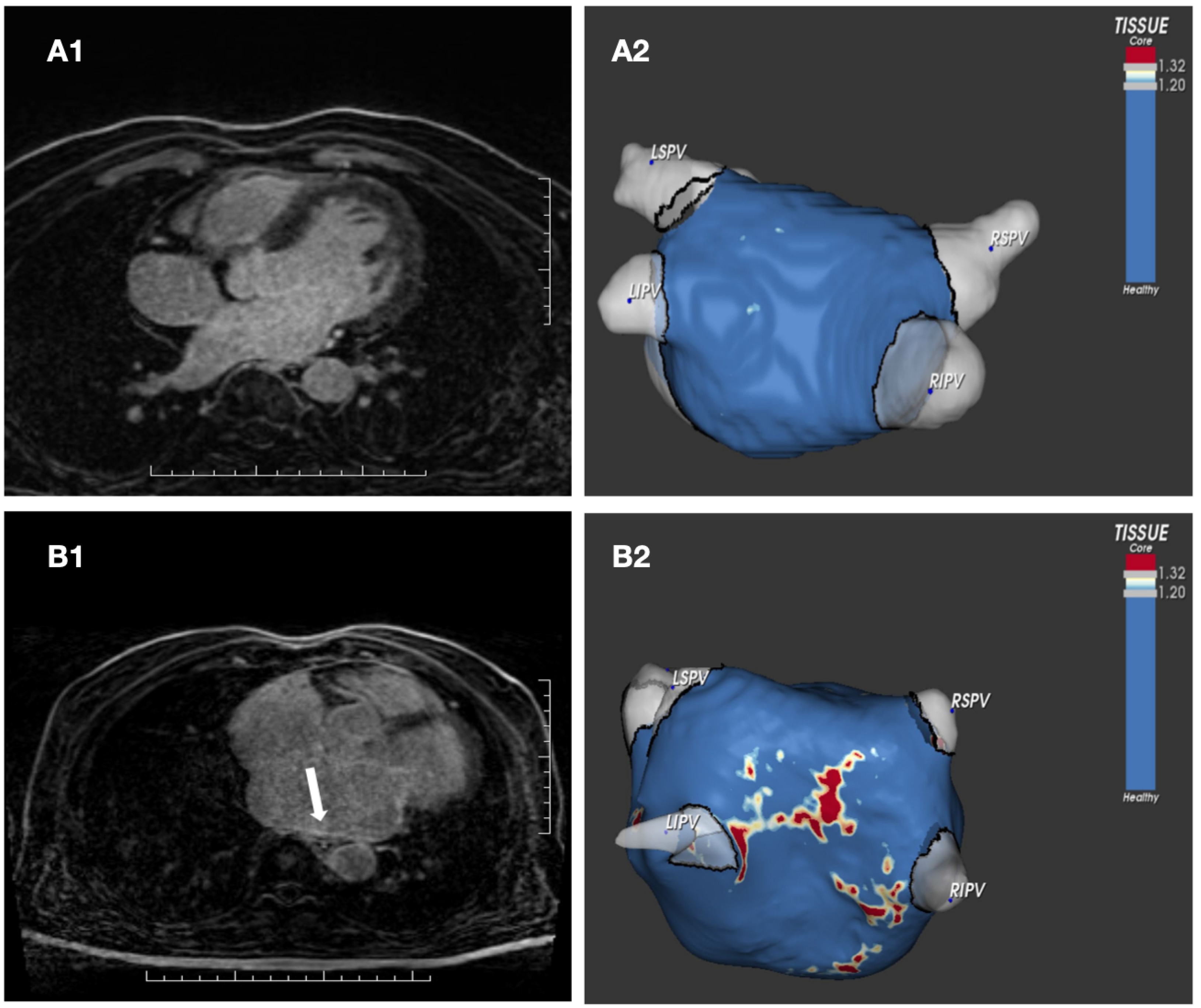
Representative images of quantitative LACI (A) and LASI (B) in a degenerative mitral regurgitation patient with atrial fibrillation. LACI, left atrioventricular coupling index; LASI, left atrial stiffness index; LAVmin, minimum left atrial volume; LVEDV, left ventricular end-diastolic volume; LASr, left atrial reservoir strain.

The LA sphericity index was defined as the ratio of the LA transverse diameter to the LA longitudinal diameter. The LACI was calculated as:

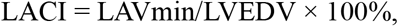

where LVEDV denotes left ventricular end-diastolic volume. The LA:LV ratio was calculated as LAVmax/LVEDV.^16^ LASI was calculated as the ratio of the average E/e’ value to LASr.^16^ Left atrial strain was assessed offline by two-dimensional speckle-tracking echocardiography using dedicated left atrial strain analysis software, following ASE/EACVI methodological recommendations. Analysis was performed using a high-quality, nonforeshortened apical four-chamber view. The left atrial endocardial border was traced, and the region of interest was manually adjusted when necessary to ensure adequate tracking throughout the cardiac cycle.^21^ Echocardiographic parameters were assessed independently by two experienced echocardiographers (X.L. and L.M.) who were blinded to CMR, biomarker, and follow-up data. Disagreements were resolved by consensus, with adjudication by a third echocardiographer (F.F.) when necessary.

### Cardiac Magnetic Resonance Imaging and LALGE Quantification

All CMR studies were performed using a 3.0-T scanner (Magnetom Vida, Siemens Healthineers, Forchheim, Germany) with retrospective electrocardiogram gating and an 8-channel cardiac coil. Cine images were acquired by balanced steady-state free precession (b-SSFP) sequence, which was used to identify the location of the left atrium. The imaging parameters were as follows: repetition time, 291.79 ms; echo time, 1.33 ms; slice thickness, 8 mm; and flip angle, 54°. Gadopentetate dimeglumine (Bayer, Germany) was administered intravenously through the antecubital vein at a dose of 0.2 mmol/kg. LGE imaging was performed 10-20 minutes after contrast administration using a free-breathing three-dimensional high-resolution inversion-recovery-prepared gradient-echo sequence. The imaging parameters were as follows: repetition time, 941.60 ms; echo time, 1.40 ms; inversion time, 350-370 ms; flip angle, 14°; and acquired voxel size, 1.25×1.25×1.25 mm. The acquisition time was approximately 10-15 minutes.

Image post-processing was performed using ADAS software (version 2.9.4, Galgo Medical S.L.). The left atrial wall contour was manually delineated and corrected on each axial slice. The software then automatically segmented the left atrial wall and blood pool and calculated the image intensity ratio (IIR) for each voxel, defined as the signal intensity of each voxel divided by the mean blood pool signal intensity. In the present study, abnormal LA wall enhancement was defined using an IIR threshold of >1.20, whereas regions with an IIR >1.32 were categorized as high-intensity enhancement.^22^ After the fibrosis threshold was defined, the IIR of each voxel was color-coded by the software: red indicated dense myocardial scar (IIR>1.32), yellow indicated interstitial fibrosis (1.20<IIR≤1.32), and blue indicated normal myocardium (IIR≤1.20). These color-coded data were then projected onto the previously reconstructed three-dimensional left atrial model to generate three-dimensional LGE-CMR images of the left atrium.

ADAS software was used to obtain parameters including left atrial wall area, LGE area, LGE percentage (**Figure 2**). LALGE percentage was calculated as follows:

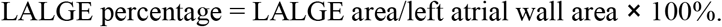

**Figure 2.**
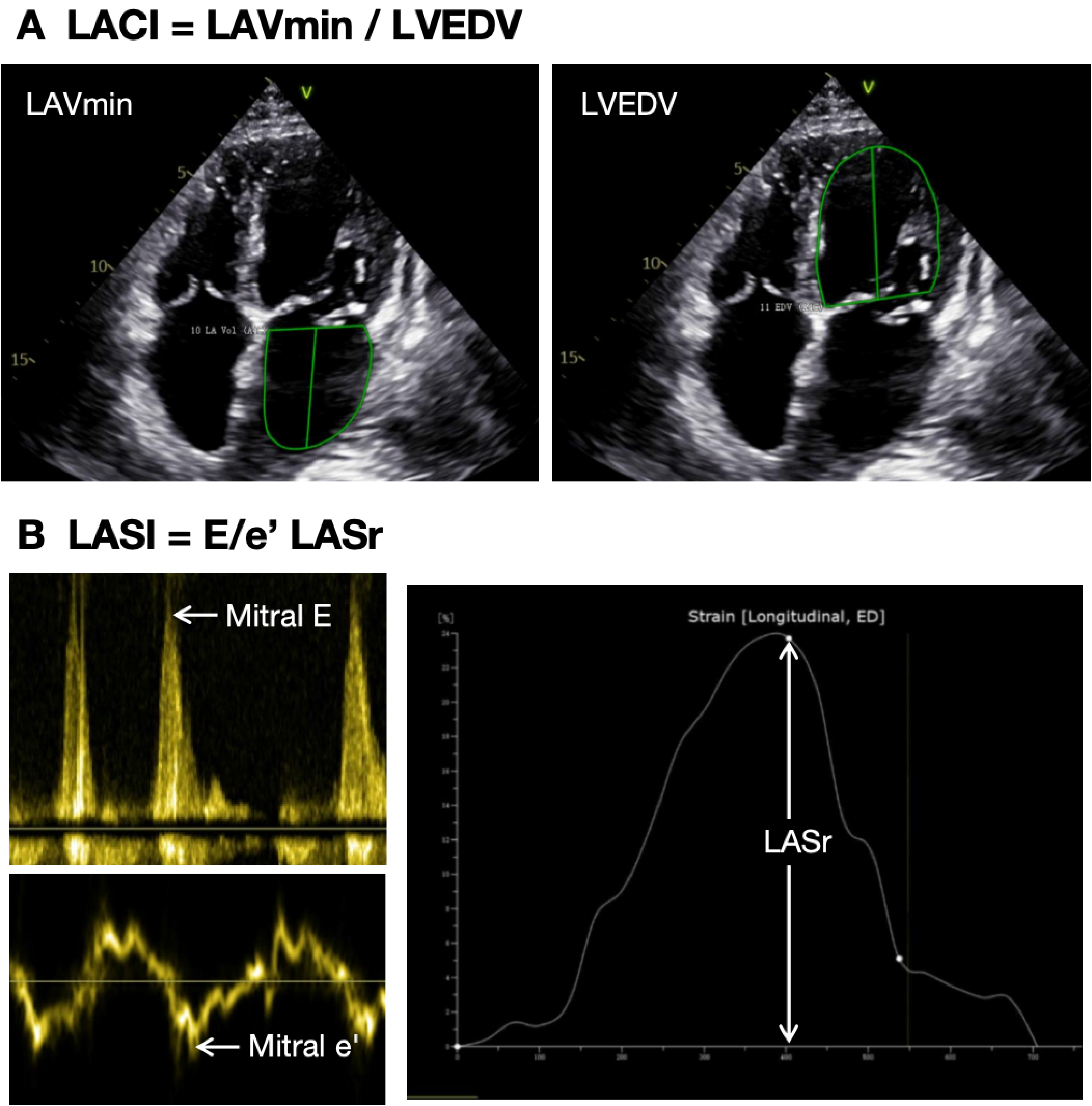
Representative cardiac magnetic resonance images and three-dimensional LGE visualizations in patients with degenerative mitral regurgitation. Panel A illustrates a patient without left atrial late gadolinium enhancement. A1 shows the acquired LGE image, and A2 shows the corresponding three-dimensional visualization after exclusion of the pulmonary veins and mitral valve region, with blue indicating normal myocardium. Panel B illustrates a patient with left atrial late gadolinium enhancement. The white arrows indicate regions of increased enhancement. B1 shows the acquired LGE image, and B2 shows the corresponding three-dimensional visualization after exclusion of the pulmonary veins and mitral valve region. Red indicates dense myocardial scar enhancement categorized as dense scar (IIR>1.32), yellow indicates interstitial fibrosis categorized as interstitial fibrosis (1.20<IIR ≤1.32), and blue indicates normal myocardium within the normal reference range (IIR ≤1.20). LGE, late gadolinium enhancement; DMR, degenerative mitral regurgitation; LSPV, left superior pulmonary vein; LIPV, left inferior pulmonary vein; RSPV, right superior pulmonary vein; RIPV, right inferior pulmonary vein; IIR, image intensity ratio.

In this study, LALGE percentage was analyzed as a standardized measure of LGE burden, and LALGE area as an absolute measure of LGE burden. In the present quantitative analysis, the pulmonary veins, mitral valve, and left atrial appendage regions were excluded from the segmentation. LALGE was assessed independently by two observers (Y.S. and L.Z.) who were blinded to the clinical and echocardiographic data. Disagreements were resolved by consensus, and any unresolved cases were adjudicated by a third experienced CMR specialist (M.L.).

### Biomarker Measurement

Fasting venous blood samples were collected at hospital admission before treatment initiation. Serum was separated by centrifugation at 3500 rpm for 15 min, aliquoted, and stored at −80°C until analysis, with repeated freeze–thaw cycles avoided. Serum C-terminal telopeptide of type I collagen (CITP; MM-50630H1; Meimian Biotechnology; China), galectin-3 (E-EL-H1470; Elabscience Biotechnology; China), and procollagen type III N-terminal propeptide (PIIINP; E-EL-H0183; Elabscience Biotechnology; China) concentrations were measured in duplicate using commercially available enzyme-linked immunosorbent assay (ELISA) kits according to the manufacturer’s instructions. Serum samples were diluted 1:10 before analysis, and measured concentrations were corrected for the dilution factor. Samples and serially diluted standards were added to pre-coated 96-well ELISA plates. Optical density was measured at 450 nm, and concentrations were calculated from the corresponding standard curves. Investigators performing the assays were blinded to AF status and imaging results. The measured biomarker levels were used to compare differences between the atrial fibrillation and non-atrial fibrillation groups.

### Statistical Analysis

Continuous variables are presented as mean ± standard deviation or median with interquartile range (IQR), as appropriate, and categorical variables as counts and percentages. Normality was assessed using the Shapiro– Wilk test. Between-group comparisons were performed using the independent-samples t test or Mann– Whitney U test for continuous variables and the chi-square test or Fisher exact test for categorical variables, as appropriate. All tests were two-sided, and *P*<0.05 was considered statistically significant.

Clinical characteristics, echocardiographic parameters, and CMR parameters were first compared between patients with and without AF. Univariable logistic regression was then performed to evaluate the associations between individual imaging parameters and AF. Because only 17 patients had AF, sensitivity analyses were performed using Firth penalized logistic regression, with each imaging parameter entered into a separate model adjusted for age and sex. Odds ratios (ORs) and 95% confidence intervals (CIs) were reported. To facilitate interpretation of the associations with cross-sectional AF status, ORs for LA sphericity index, LA:LV, and LASI were reported per 0.1-unit increase, whereas the OR for LACI was reported per 10-percentage-point increase. Other ORs were expressed per one unit of the corresponding measurement. Receiver operating characteristic (ROC) curve analysis was used to evaluate the discriminative performance of individual and combined imaging parameters. Area under the curves (AUCs) and corresponding 95% CI were calculated. Optimal cutoff values for individual and combined imaging parameters were determined using the Youden index, and sensitivity, specificity, positive predictive value (PPV), and negative predictive value (NPV) were calculated at the corresponding cutoff values. Pairwise AUC comparisons were performed using the DeLong test, with Holm adjustment for multiple comparisons.

Variance inflation factors (VIFs) were used to assess potential multicollinearity among predictors included in multiparameter models. Given the limited sample size and number of AF events in this study, model complexity was restricted to two predictors. LASr and LACI were selected to represent LA mechanical function and atrioventricular coupling, respectively, whereas LALGE area and LALGE percentage were selected to represent absolute and normalized enhancement burden. Selection was also supported by the univariable and ROC findings. The resulting models were considered exploratory and were not externally validated. Spearman rank correlation coefficients were used to assess the relationships between serum CITP levels and imaging parameters. All statistical analyses were performed using RStudio with R version 4.4.2.

## Results

### Study Population

Among 48 DMR patients who underwent echocardiography and CMR, 2 were excluded because of inadequate CMR image quality. A total of 46 patients were included in the final analysis, and the baseline characteristics of the study population are presented in **Table 1**. The median age was 62.0 years (IQR, 55.00, 70.75 years), 33 patients (71.7%) were men, mean body mass index (BMI) was 24.68 ± 3.48 kg/m², and 17 (37.0%) had atrial fibrillation, including 14 with persistent AF and 3 with paroxysmal AF. 14 (30.4%) underwent transcatheter edge-to-edge repair, 22 (47.8%) underwent surgical mitral valve repair, 9 (19.6%) underwent mitral valve replacement, and 1 (2.2%) underwent no mitral valve intervention. Compared with patients without AF, those with AF were older (*P*=0.005), were more likely to have pulmonary hypertension (*P*<0.001), and had lower left ventricular ejection fraction (*P*=0.023). Patients with AF had higher levels of N-terminal pro-B-type natriuretic peptide (NT-proBNP; *P*<0.001), lower lipoprotein(a) (*P*=0.027) and lower estimated glomerular filtration rate (eGFR; *P*=0.006). No significant between-group differences were observed in sex, BMI, major comorbidities, MR severity, or left ventricular dimensions and volumes.

**Table 1.** Baseline Characteristics of Patients With Degenerative Mitral Regurgitation.

| **Variables** | **All (N=46)** | **AF group (n=17)** | **Non-AF group (n=29)** | **P Value** |
| --- | --- | --- | --- | --- |
| Male | 33 (71.7%) | 12 (70.6%) | 21 (72.4%) | 1.000 |
| Age, years | 62.00 (55.00, 70.75) | 70.00 (62.00, 76.00) | 61.00 (51.00, 68.00) | 0.005 |
| BMI, kg/m<sup>2</sup> | 24.68 ± 3.48 | 24.31 ± 4.23 | 24.89 ± 3.02 | 0.622 |
| **Comorbidity** |  |  |  |  |
| Hypertension | 24 (52.2%) | 9 (52.9%) | 15 (51.7%) | 1.000 |
| Diabetes mellitus | 9 (19.6%) | 6 (35.3%) | 3 (10.3%) | 0.058 |
| Chronic obstructive pulmonary disease | 2 (4.3%) | 1 (5.9%) | 1 (3.4%) | 1.000 |
| Stroke | 2 (4.3%) | 0 (0.0%) | 2 (6.9%) | 0.524 |
| Coronary artery disease | 7 (15.2%) | 4 (23.5%) | 3 (10.3%) | 0.397 |
| Prior coronary intervention or bypass surgery | 5 (10.9%) | 2 (11.8%) | 3 (10.3%) | 1.000 |
| **AF and treatment-related characteristics** |  |  |  |  |
| Persistent AF | 14 (30.4%) | 14 (82.4%) | 0 (0.0%) | NA |
| Paroxysmal AF | 3 (6.5%) | 3 (17.6%) | 0 (0.0%) | NA |
| Mitral valve intervention strategy |  |  |  | 0.004 |
| Transcatheter edge-to-edge mitral repair | 14 (30.4%) | 9 (52.9%) | 5 (17.2%) |  |
| Surgical mitral valve repair | 22 (47.8%) | 3 (17.6%) | 19 (65.5%) |  |
| Surgical mitral valve replacement | 9 (19.6%) | 5 (29.4%) | 4 (13.8%) |  |
| No mitral valve intervention | 1 (2.2%) | 0 (0.0%) | 1 (3.4%) |  |
| **Echocardiographic parameters** |  |  |  |  |
| Pulmonary hypertension | 17 (37.0%) | 12 (70.6%) | 5 (17.2%) | <0.001 |
| Tricuspid regurgitation | 29 (63.0%) | 11 (64.7%) | 18 (62.1%) | 1.000 |
| Left ventricular ejection fraction, % | 67.09 ± 5.88 | 64.41 ± 6.08 | 68.66 ± 5.25 | 0.023 |
| Left ventricular end-diastolic diameter, mm | 56.98 ± 5.41 | 56.41 ± 6.01 | 57.31 ± 5.10 | 0.609 |
| Left ventricular end-diastolic volume, mL | 124.62 (101.61, 139.39) | 107.28 (99.86, 127.06) | 127.04 (118.53, 143.75) | 0.080 |
| MR severity grade |  |  |  |  |
| Moderate-to-severe | 7 (15.2%) | 3 (17.6%) | 4 (13.8%) | 1.000 |
| Severe | 39 (84.8%) | 14 (82.4%) | 25 (86.2%) | 1.000 |
| Left atrial echocardiographic parameters |  |  |  |  |
| LA transverse diameter, mm | 50.04 ± 10.33 | 52.68 ± 14.58 | 48.50 ± 6.59 | 0.277 |
| LA longitudinal diameter, mm | 59.38 ± 10.11 | 65.23 ± 11.95 | 55.95 ± 7.04 | 0.008 |
| LA sphericity index | 0.85 ± 0.15 | 0.80 ± 0.15 | 0.88 ± 0.14 | 0.115 |
| LAVmax, mL | 85.44 (68.50, 138.90) | 138.62 (83.36, 159.76) | 75.89 (68.48, 94.57) | 0.032 |
| LAVmin, mL | 48.84 (39.80, 84.23) | 102.54 (60.43, 131.75) | 43.65 (34.56, 55.06) | <0.001 |
| LAEF, % | 37.21 ± 16.77 | 23.78 ± 12.64 | 45.09 ± 13.67 | <0.001 |
| LASr, % | 25.46 ± 10.69 | 17.47 ± 8.72 | 30.14 ± 8.87 | <0.001 |
| LAScd, % | -17.80 ± 7.98 | -13.72 ± 7.56 | -20.19 ± 7.33 | 0.008 |
| LACI, % | 39.98 (30.76, 69.79) | 73.90 (61.33, 123.89) | 35.05 (28.05, 45.60) | <0.001 |
| LA: LV | 0.72 (0.57, 1.01) | 1.00 (0.81, 1.44) | 0.64 (0.53, 0.88) | 0.005 |
| LASI | 0.44 (0.30, 0.63) | 0.64 (0.51, 1.24) | 0.34 (0.28, 0.46) | <0.001 |
| E/e' | 10.53 (8.20, 11.97) | 11.25 (9.67, 17.20) | 10.40 (8.11, 11.33) | 0.280 |
| CMR LGE parameters |  |  |  |  |
| Total LA wall area, cm<sup>2</sup> | 128.00 (101.03, 174.47) | 179.41 (157.88, 216.78) | 117.67 (96.84, 130.53) | <0.001 |
| LALGE area, cm<sup>2</sup> | 9.04 ± 6.94 | 13.89 ± 8.27 | 6.20 ± 3.97 | 0.002 |
| LALGE percentage, % | 6.35 ± 4.17 | 8.16 ± 4.92 | 5.29 ± 3.30 | 0.043 |
| Laboratory values |  |  |  |  |
| Hemoglobin, g/L | 146.63 ± 14.56 | 145.00 ± 18.05 | 147.59 ± 12.32 | 0.605 |
| Creatinine, µmol/L | 79.16 ± 19.10 | 86.36 ± 22.60 | 74.94 ± 15.64 | 0.077 |
| eGFR, mL/min/1.73 m<sup>2</sup> | 85.12 ± 18.82 | 74.70 ± 20.13 | 91.44 ± 15.09 | 0.006 |
| LDL cholesterol, mmol/L | 2.64 ± 0.94 | 2.44 ± 0.74 | 2.75 ± 1.04 | 0.241 |
| Lipoprotein (a), g/L | 1.50 ± 0.32 | 1.37 ± 0.30 | 1.58 ± 0.31 | 0.027 |
| NT-proBNP, pg/mL | 217.00 (70.00, 1268.00) | 1303.00 (649.00, 2541.00) | 88.55 (57.67, 220.75) | <0.001 |
| D-dimer, µg/mL | 0.22 (0.13, 0.43) | 0.26 (0.13, 0.68) | 0.21 (0.13, 0.33) | 0.351 |

AF, atrial fibrillation; BMI, body mass index; MR, mitral regurgitation; LA, left atrial; LAVmax, maximum left atrial volume; LAVmin, minimum left atrial volume; LAEF, left atrial ejection fraction; LASr, left atrial reservoir strain; LAScd, left atrial conduit strain; LACI, left atrioventricular coupling index; LA:LV, left atrial-to-left ventricular volume ratio; LASI, left atrial stiffness index; CMR, cardiac magnetic resonance; LALGE, left atrial late gadolinium enhancement; eGFR, estimated glomerular filtration rate; LDL, low-density lipoprotein; NT-proBNP, N-terminal pro-B-type natriuretic peptide; NA, not applicable.

Follow-up data were available for 45 patients (97.8%), whereas 1 patient (2.2%) was lost to follow-up. During a median follow-up of 9.2 months (IQR, 7.7-12.2 months) among patients with available follow-up, one patient in the AF group who had undergone transcatheter edge-to-edge repair subsequently underwent catheter ablation for AF at 8 months, and another patient with prior transcatheter edge-to-edge repair was hospitalized for heart failure at 8 months. In the non-AF group, recurrent MR was observed at 8 and 7 months of follow-up, respectively, in two patients who had undergone surgical mitral valve repair. No deaths, strokes, or other prespecified events were observed.

### Echocardiographic and CMR Findings

Representative images of quantitative LACI (A) and LASI (B) in a DMR patient with atrial fibrillation are shown in **Figure 1**. Compared with patients without AF, those with AF had a greater LA longitudinal diameter (*P*=0.008) and larger maximum and minimum LA volumes (*P*=0.032 and *P*<0.001, respectively), whereas LA transverse diameter and LA sphericity index did not differ significantly between groups. LA mechanical function was also more impaired in the patients with AF, as indicated by lower LA ejection fraction (*P*<0.001), lower LASr (*P*<0.001), and a lower absolute magnitude of LAScd (*P*=0.008). In addition, patients with AF had higher LACI (*P*<0.001), LA:LV (*P*=0.005), and LASI (*P*<0.001) values.

Representative CMR images of LALGE are shown in **Figure 2**. Patients with AF had a larger total LA wall area than those without AF (*P*<0.001). Both the absolute and normalized burdens of LALGE were also greater in the AF group, as reflected by higher LALGE area (*P*=0.002) and LALGE percentage (*P*=0.043). These findings indicated a more advanced LA remodeling phenotype in patients with AF, characterized by LA enlargement, impaired mechanical function, abnormal atrioventricular coupling, and a greater CMR-detected atrial enhancement burden.

### Associations of Echocardiographic and CMR Parameters With Atrial Fibrillation

In univariable logistic regression analyses, multiple echocardiographic and CMR indices of LA remodeling were associated with prevalent AF (**Table 2**). In age- and sex-adjusted sensitivity analyses using Firth penalized logistic regression, these associations remained statistically significant. Larger LAVmax (adjusted OR, 1.02; 95% CI, 1.01-1.04; *P*=0.006), larger LAVmin (adjusted OR, 1.03; 95% CI, 1.01-1.06; *P*<0.001), higher LACI (adjusted OR per 10-percentage-point increase, 1.48; 95% CI, 1.18-2.23; *P*<0.001), and less negative LAScd (adjusted OR, 1.11; 95% CI, 1.01-1.24; *P*=0.026) were associated with higher odds of AF. In contrast, higher LAEF (adjusted OR, 0.91; 95% CI, 0.84-0.96; *P*<0.001) and higher LASr (adjusted OR, 0.86; 95% CI, 0.76-0.94; *P*<0.001) were associated with lower odds of AF. Each 0.1-unit increase in LA:LV and LASI was associated with higher odds of AF, with adjusted ORs of 1.28 (95% CI, 1.08-1.59; *P*=0.004) and 1.31 (95% CI, 1.07-1.72; *P*=0.002), respectively. Among the CMR-derived parameters, greater total LA wall area (adjusted OR, 1.04; 95% CI, 1.02-1.06; *P*<0.001), larger LALGE area (adjusted OR, 1.20; 95% CI, 1.07-1.41; *P*<0.001), and higher LALGE percentage (adjusted OR, 1.19; 95% CI, 1.02-1.42; *P*=0.025) remained associated with higher odds of AF.

**Table 2.** Associations Between Imaging Parameters and Atrial Fibrillation in Degenerative Mitral Regurgitation.

| Variables | Unadjusted OR (95% CI) | P Value | Adjusted OR (95% CI)* | P Value |
| --- | --- | --- | --- | --- |
| **Echocardiographic parameters** |  |  |  |  |
| LA sphericity index<sup>†</sup> | 0.69 (0.42-1.06) | 0.114 | 0.72 (0.44-1.10) | 0.133 |
| LAVmax, mL | 1.02 (1.00-1.03) | 0.012 | 1.02 (1.01-1.04) | 0.006 |
| LAVmin, mL | 1.04 (1.01-1.06) | 0.002 | 1.03 (1.01-1.06) | <0.001 |
| LAEF, % | 0.90 (0.85-0.95) | <0.001 | 0.91 (0.84-0.96) | <0.001 |
| LASr, % | 0.85 (0.77-0.93) | <0.001 | 0.86 (0.76-0.94) | <0.001 |
| LAScd, % | 1.13 (1.03-1.25) | 0.013 | 1.11 (1.01-1.24) | 0.026 |
| LACI, %<sup>†</sup> | 1.79 (1.32-2.76) | 0.002 | 1.48 (1.18-2.23) | <0.001 |
| LA:LV<sup>†</sup> | 1.38 (1.13-1.79) | 0.005 | 1.28 (1.08-1.59) | 0.004 |
| LASI<sup>†</sup> | 1.45 (1.16-2.02) | 0.007 | 1.31 (1.07-1.72) | 0.002 |
| **CMR LGE parameters** |  |  |  |  |
| Total LA wall area, cm<sup>2</sup> | 1.04 (1.02-1.06) | <0.001 | 1.04 (1.02-1.06) | <0.001 |
| LALGE area, cm<sup>2</sup> | 1.23 (1.10-1.43) | 0.002 | 1.20 (1.07-1.41) | <0.001 |
| LALGE percentage, % | 1.20 (1.03-1.44) | 0.032 | 1.19 (1.02-1.42) | 0.025 |

OR, odds ratio; CI, confidence interval; LA, left atrium; LAVmax, maximum left atrial volume; LAVmin, minimum left atrial volume; LAEF, left atrial ejection fraction; LASr, left atrial reservoir strain; LAScd, left atrial conduit strain; LACI, left atrioventricular coupling index; LA:LV, left atrial- to-left ventricular volume ratio; LASI, left atrial stiffness index; CMR, cardiac magnetic resonance; LALGE, left atrial late gadolinium enhancement. \*Age- and sex-adjusted ORs, 95% CIs, and P values were estimated using Firth penalized logistic regression, with each imaging parameter entered into a separate model. <sup>†</sup> ORs for LA sphericity index, LA:LV, and LASI are reported per 0.1-unit increase; the OR for LACI is reported per 10-percentage-point increase.

### Echocardiographic, CMR, and Multimodality Models

Among the four selected individual imaging parameters, LACI had the highest observed AUC (0.85; 95% CI, 0.72-0.99), followed by LASr (AUC, 0.84; 95% CI, 0.72-0.96), LALGE area (AUC, 0.78; 95% CI, 0.61-0.94), and LALGE percentage (AUC, 0.68; 95% CI, 0.50-0.86). The corresponding exploratory cutoff values were 58.59%, 24.65%, 13.73 cm², and 9.63%, respectively. At its optimal cutoff, LACI yielded a sensitivity of 82.4% and a specificity of 86.2%, whereas LALGE area yielded a specificity of 96.6% and a PPV of 90.9% at its corresponding cutoff (**Table 3)**.

**Table 3.**
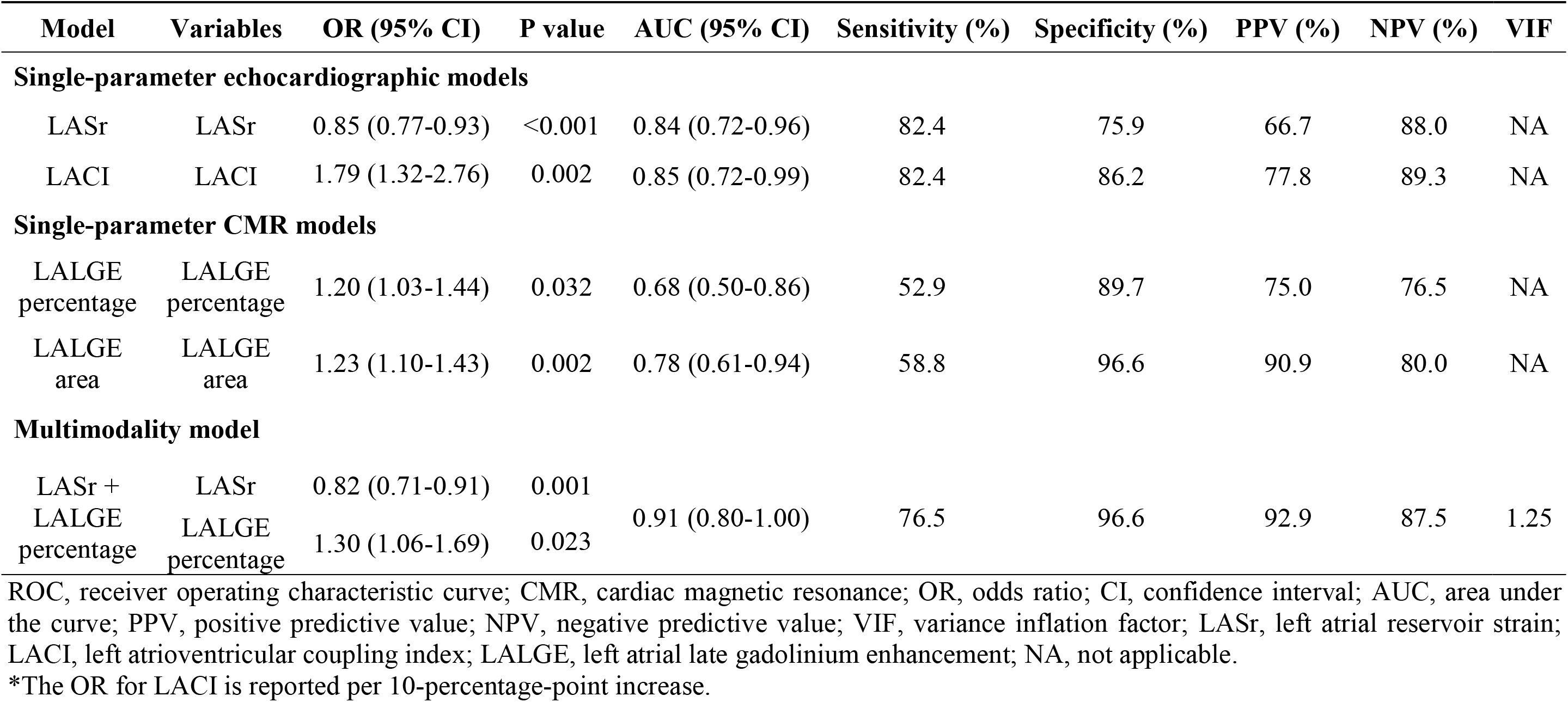
Logistic Regression Analysis and ROC Performance of Echocardiographic, CMR, and Multimodality Models for Identifying Atrial Fibrillation.

The multiparameter echocardiographic model combining LASr and LACI achieved an AUC of 0.87 (95% CI, 0.74-1.00), with a sensitivity of 76.5%, specificity of 96.6%, PPV of 92.9%, and NPV of 87.5%. Its AUC was numerically higher than those of LASr and LACI alone, which were 0.84 and 0.85, respectively. The multiparameter CMR model combining LALGE percentage and LALGE area yielded an AUC of 0.81 (95% CI, 0.66-0.95), with a sensitivity of 76.5%, specificity of 82.8%, PPV of 72.2%, and NPV of 85.7%. Its AUC was numerically higher than those of LALGE percentage and LALGE area alone, which were 0.68 and 0.78, respectively (**Supplementary Table 1**).

Among the multimodality models, the combination of LASr and LALGE percentage showed the highest observed AUC of 0.91 (95% CI, 0.80-1.00), with a sensitivity of 76.5%, specificity of 96.6%, PPV of 92.9%, and NPV of 87.5% (**Figure 3**). ROC curves of all models were shown in **Supplementary Figure 1**. The model combining LASr and LALGE area yielded an AUC of 0.90 (95% CI, 0.79-1.00), with a sensitivity of 82.4%, specificity of 96.6%, PPV of 93.3%, and NPV of 90.3%. Models combining LACI with LALGE percentage or LALGE area yielded AUCs of 0.89 and 0.89, respectively. VIF values ranged from 1.00 to 1.25 across the multimodality models, indicating no evidence of relevant multicollinearity. However, pairwise DeLong tests showed no statistically significant differences in AUC among the models after Holm adjustment for multiple comparisons (all adjusted *P*≥0.188).

**Figure 3.**
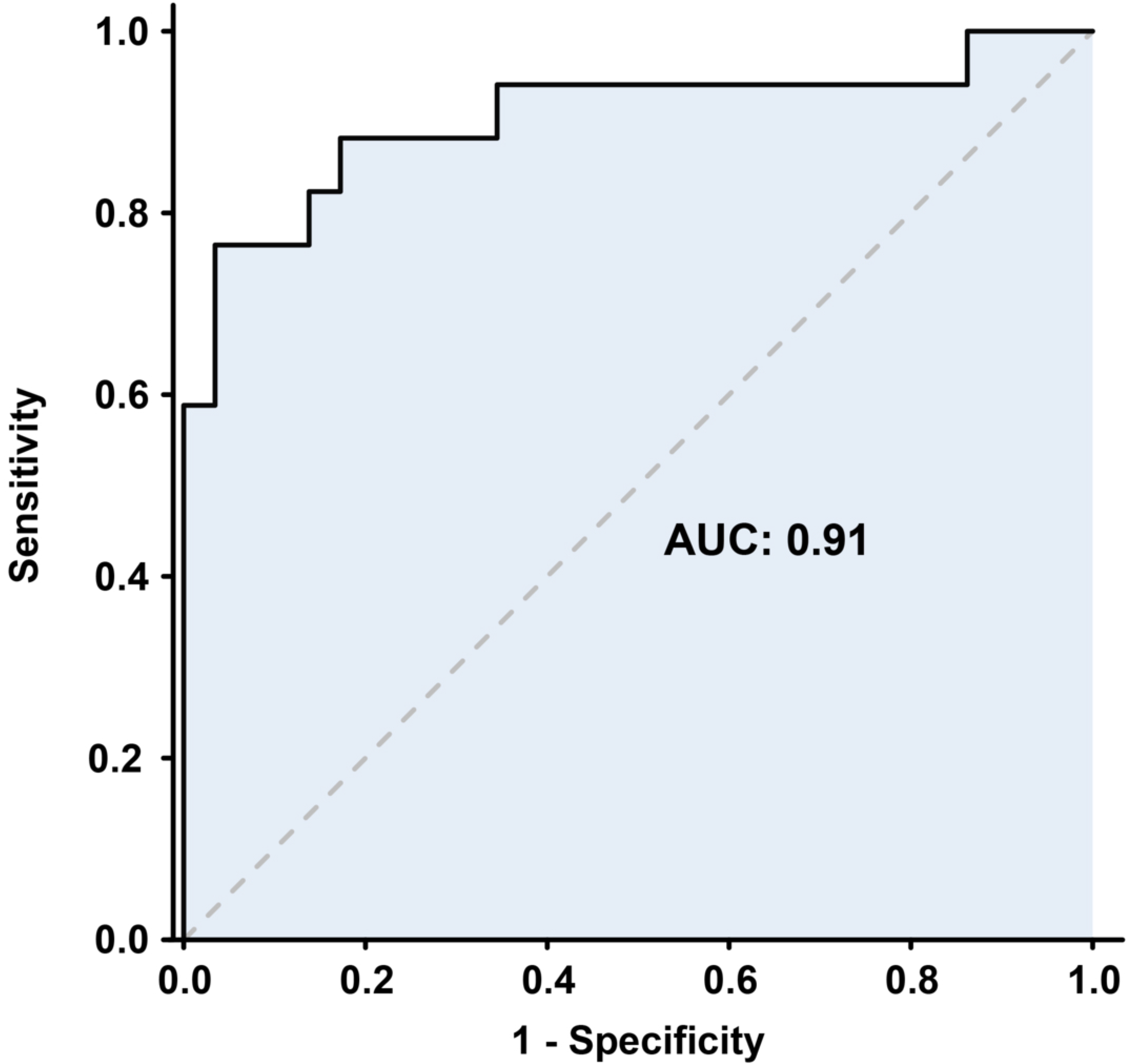
ROC curve of the LASr + LALGE percentage model for identifying atrial fibrillation The ROC curve for the model combining LASr and LALGE percentage is shown, with an AUC of 0.91 (95% CI, 0.80-1.00). ROC, receiver operating characteristic; LASr, left atrial reservoir strain; LALGE, left atrial late gadolinium enhancement; AUC, area under the curve; CI, confidence interval.

### ELISA Analysis

In the exploratory ELISA-based biomarker analysis (**Table 4** and **Figure 4**), serum CITP levels were higher in patients with AF than in those without AF (2.99±1.65 vs. 1.71±1.07 ng/mL, *P*=0.009). Galectin-3 and PIIINP were also higher in the AF group, but the between-group differences were not statistically significant (*P*=0.172 and *P*=0.249, respectively). In exploratory Spearman correlation analyses, serum CITP levels were not significantly correlated with LASr, LACI, LALGE area, or LALGE percentage (all *P*>0.05; **Supplementary Table 2**).

**Figure 4.**
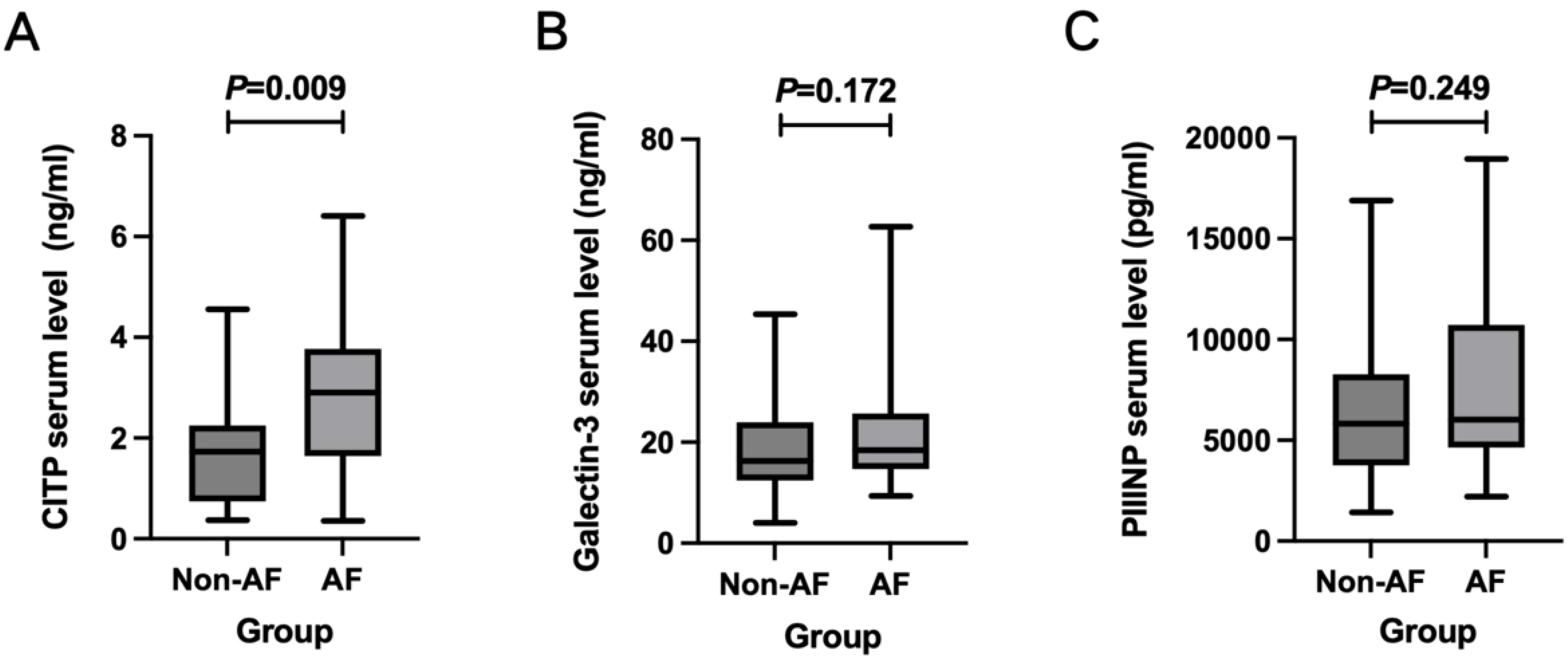
Serum fibrosis-related biomarker levels in patients with degenerative mitral regurgitation by ELISA. Panels A, B, and C show serum CITP, galectin-3, and PIIINP levels, respectively. *P* < 0.05 was considered statistically significant. ELISA, enzyme-linked immunosorbent assay; CITP, C-terminal telopeptide of type I collagen; PIIINP, procollagen type III N-terminal propeptide.

**Table 4.** Serum Fibrosis-Related Biomarker Levels in Patients With Degenerative Mitral Regurgitation by ELISA.

| **Variables** | **All (N=46)** | **AF group (n=17)** | **Non-AF group (n=29)** | **P Value** |
| --- | --- | --- | --- | --- |
| CITP, ng/mL | 1.89 (1.23, 2.94) | 2.99 ± 1.65 | 1.71 ± 1.07 | 0.009 |
| Galectin-3, ng/mL | 16.93 (13.23, 25.61) | 18.43 (14.82, 25.63) | 16.27 (12.84, 22.07) | 0.172 |
| PIIINP, pg/mL | 5910.43 (4070.28, 8288.33) | 6026.19 (4701.70, 10367.61) | 5824.30 (3793.03, 8227.07) | 0.249 |

AF, atrial fibrillation; ELISA, enzyme-linked immunosorbent assay; CITP, C-terminal telopeptide of type I collagen; PIIINP, procollagen type III N-terminal propeptide.

## Discussion

The principal findings of this study are as follows. First, patients with isolated DMR and AF exhibited more advanced LA structural remodeling, greater impairment of LA function, abnormal left atrioventricular coupling, and a higher CMR-derived LALGE burden than patients without AF. Second, echocardiographic measures of LA function, particularly LASr and LACI, and CMR-derived measures of LALGE were associated with AF. Finally, the combination of LASr and LALGE percentage yielded the highest observed AUC among the evaluated models, suggesting that these modalities may provide complementary information by capturing distinct functional and tissue characteristics. Few studies have integrated echocardiographic measures of LA function with CMR-derived LALGE measures in patients with isolated DMR, and the present findings provide a multidimensional description of AF-associated atrial remodeling in this setting.

Chronic regurgitant volume overload in DMR may lead to progressive LA dilatation, increased wall stress, mechanical dysfunction, and alterations in atrial myocardial composition.^23,24^ These structural and functional changes may create a substrate that facilitates AF, whereas AF itself may further impair atrial mechanical function and promote adverse remodeling.^11,24^ Accordingly, the association between AF and more advanced LA remodeling observed in this study should be interpreted as reflecting a bidirectional interaction rather than a causal relationship.

Echocardiographic assessment provides accessible measures of LA size and mechanical function. The present study aimed to further explore the potential clinical value of these parameters in patients with DMR. Previous studies have shown that impaired LASr in patients with severe MR is associated with adverse clinical outcomes.^25^ Moreover, although LA strain reflects atrial mechanical function, its relationship with histologically assessed atrial fibrosis has not been consistent across studies.^26^ Previous studies have shown that LACI provides independent and incremental prognostic value for heart failure, atrial fibrillation, and mortality, outperforming isolated atrial or ventricular measures and supporting its use as a marker of dynamic remodeling and cardiovascular risk stratification.^27,28^ In the present study, LASr and LACI showed similar single-parameter discrimination for AF, suggesting that impaired reservoir function and abnormal atrioventricular volumetric coupling represent related but distinct features of atrial remodeling. When LASr and LACI were included in the same model, the association of LASr was attenuated, whereas LACI remained statistically significant. Given the limited number of AF events and the resulting uncertainty in model estimates, this finding should be interpreted cautiously and should not be considered evidence that LACI is superior to LASr.

Previous LALGE studies have primarily focused on general populations with AF or on patients undergoing catheter ablation, with particular emphasis on methods for left atrial wall segmentation and fibrosis quantification. Existing imaging studies have also highlighted that, because the left atrial wall is extremely thin, LGE quantification is susceptible to variations in spatial resolution, image quality, segmentation approaches, and thresholding methods. In recent years, three-dimensional LGE-CMR has increasingly been used for the noninvasive assessment of LA tissue characteristics.^29,30^ Conventional two-dimensional LGE-CMR has relatively limited spatial resolution and typically uses anisotropic voxels, resulting in substantial imaging errors when assessing the thin atrial wall. In the present study, an isotropic spatial resolution of 1.25×1.25×1.25 mm was achieved, enabling quantitative assessment and three-dimensional visualization of the spatial distribution of IIR-defined LA wall enhancement. This technique may provide one of the most suitable currently available noninvasive imaging approaches for detecting and characterizing atrial myocardial fibrosis.

Echocardiography and CMR assess different dimensions of LA remodeling. Echocardiographic strain reflects atrial mechanical function, left atrial volume characterizes structural remodeling, and LGE-CMR provides information on the underlying tissue substrate. Accordingly, no single imaging dimension can fully characterize the complex phenotype of atrial myopathy.^31^ The multimodality models integrated LASr as a marker of mechanical function, LACI as an index of left atrioventricular volumetric coupling, and LALGE area and LALGE percentage as measures of the absolute and normalized tissue enhancement burden, respectively.^22,32,33^ The low VIF values observed in the multimodality models indicated no substantial multicollinearity among the included predictors, and the higher AUCs of the combined models may suggest potential complementary value. Although a broad spectrum of left atrial remodeling is observed in patients with MR, few studies have systematically assessed left atrial structure, mechanical function, and imaging surrogates of the atrial tissue substrate in patients with DMR. The overall aim was to explore a noninvasive approach for characterizing atrial remodeling by comparing multidimensional imaging features in this population. In addition, longitudinal follow-up of this cohort is ongoing to further determine the potential value of these imaging markers in optimizing surgical strategies and guiding long-term prognostic management.

The exploratory biomarker analysis provided additional, although limited, information on systemic collagen turnover. Serum CITP levels were higher in patients with AF, but CITP was not significantly correlated with LASr, LACI, LALGE area, or LALGE percentage. CITP reflects systemic type I collagen turnover rather than left atrium-specific fibrosis, whereas LALGE represents a localized imaging phenotype.^34,35^ Collagen turnover activity may not necessarily parallel the burden of established fibrosis, and the findings may also be influenced by renal function, age, limited sample size, and biological variability. Thus, the biomarker findings should be regarded as hypothesis-generating and do not establish a direct relationship between circulating biomarkers and localized imaging-defined atrial fibrosis.

In summary, the present study has several notable strengths. First, the inclusion of a relatively homogeneous cohort of patients with isolated DMR, together with the exclusion of cardiomyopathies and other concomitant left-sided valvular diseases, reduced potential confounding from alternative causes of left atrial remodeling. In addition, the study combined quantitative echocardiographic measures of LA structure and function with high-resolution three-dimensional LGE-CMR assessment of atrial enhancement. This multidimensional approach allowed structural, mechanical, coupling, and tissue-enhancement features of AF-associated atrial remodeling to be evaluated within the same cohort.

### Limitations

Several limitations should be acknowledged. First, this was a single-center retrospective study with a relatively small sample size and limited number of AF events, which may reduce the precision of the regression estimates and increase the risk of model overfitting. Second, the primary analyses were cross-sectional, and the relatively short follow-up duration and small number of clinical events precluded meaningful prognostic analyses. Third, LALGE quantification may be influenced by image quality, spatial resolution, segmentation strategy, and thresholding methods, and histological validation was not available. Fourth, although age-and sex-adjusted sensitivity analyses were performed using Firth penalized logistic regression, residual confounding by other variables cannot be excluded because of the limited number of AF events. Finally, the multimodality models and optimal cutoff values were derived from the same cohort without internal or external validation and should therefore be considered exploratory. Future research should aim to validate these findings in larger, multicenter cohorts, and explore whether LALGE can be integrated into clinical decision-making algorithms for AF surveillance and management in DMR, including early rhythm control strategies and candidacy for catheter ablation or mitral intervention.

## Conclusions

In patients with isolated DMR, AF was associated with a more advanced LA remodeling phenotype, characterized by LA enlargement, impaired mechanical function, abnormal left atrioventricular coupling, and a greater CMR-derived LALGE burden. Echocardiographic LA functional indices and CMR-derived LALGE measures may provide complementary information for the discrimination of AF. These exploratory findings require validation in larger prospective cohorts.

## IRB information

This retrospective study was approved by the ethics committee of Fuwai Hospital, Chinese Academy of Medical Sciences (No. 2025-2621).

## Disclosures

The authors have no conflicts of interest to declare.

## Data Availability

The datasets generated and/or analyzed during the current study are not publicly available because they contain potentially identifiable clinical and imaging data and are subject to institutional ethical and data-protection requirements. De-identified data may be made available by the corresponding author upon reasonable request.

## Acknowledgement

None.

## Funding

This work was supported by Chinese Academy of Medical Sciences, CAMS Innovation Fund for Medical Sciences (2024-I2M-C&T-C-006); Chinese Academy of Medical Sciences, CAMS Innovation Fund for Medical Sciences (2023-I2M-C&T-B-050).

## Author contributions

Conceptualization: Fang Fang, Minjie Lu, Xiangbin Pan; Methodology: Xinmu Li, Yanyan Song, Yiran Hu; Formal analysis: Xinmu Li; Investigation: Xinmu Li, Yanyan Song; Data curation: Xinmu Li, Yanyan Song, Yiran Hu; Visualization: Xinmu Li, Yanyan Song, Yiran Hu; Writing-original draft: Xinmu Li, Yanyan Song, Yiran Hu.; Writing-review & editing: all authors; Supervision.; Funding acquisition: Fang Fang, Minjie Lu, Xiangbin Pan

## Supplementary materials

**Supplementary Figure 1. ROC curves of all evaluated imaging models for identifying atrial fibrillation.** Each colored curve represents a different model, with the corresponding model name and AUC shown in the figure legend. ROC, receiver operating characteristic curve; AUC, area under the curve; LASr, left atrial reservoir strain; LACI, left atrioventricular coupling index; LALGE, left atrial late gadolinium enhancement.

**Supplementary Table 1.** Logistic Regression and ROC Performance of Multiparameter Imaging Models for Identifying Atrial Fibrillation

**Supplementary Table 2.** Spearman Correlations Between Serum CITP Levels and Left Atrial Imaging Parameters

## Notes

### Competing Interest Statement

The authors have declared no competing interest.

### Clinical Trial

This was a single-center retrospective cross-sectional observational study based on existing clinical and imaging data. And therefore the study was not registered in a clinical trial registry.

### Author Declarations

The study was conducted in accordance with the Declaration of Helsinki and approved by the Ethics Committee of Fuwai Hospital, Chinese Academy of Medical Sciences (No. 2025-2621).

## References

1. Coffey S, Roberts-Thomson R, Brown A, Carapetis J, Chen M, Enriquez-Sarano M, Zühlke L, Prendergast BD. Global epidemiology of valvular heart disease. Nat Rev Cardiol. 2021;18:853–864.

2. Wunderlich NC, Beigel R, Rader F, Franke J, Siegel RJ. Degenerative Mitral Regurgitation: Assessment, Physical Examination, and Imaging. Curr Cardiol Rep. 2019;21:85.

3. Deferm S, Bertrand PB, Verbrugge FH, Verhaert D, Rega F, Thomas JD, Vandervoort PM. Atrial Functional Mitral Regurgitation: JACC Review Topic of the Week. J Am Coll Cardiol. 2019;73:2465–2476.

4. Delgado V, Ajmone Marsan N, Bonow RO, Hahn RT, Norris RA, Zühlke L, Borger MA. Degenerative mitral regurgitation. Nat Rev Dis Primers. 2023;9:70.

5. Chugh SS, Havmoeller R, Narayanan K, Singh D, Rienstra M, Benjamin EJ, Gillum RF, Kim YH, McAnulty JH, Jr., Zheng ZJ, et al. Worldwide epidemiology of atrial fibrillation: a Global Burden of Disease 2010 Study. Circulation. 2014;129:837–847.

6. Ko D, Chung MK, Evans PT, Benjamin EJ, Helm RH. Atrial Fibrillation: A Review. Jama. 2025;333:329–342.

7. Benjamin EJ, Wolf PA, D’Agostino RB, Silbershatz H, Kannel WB, Levy D. Impact of atrial fibrillation on the risk of death: the Framingham Heart Study. Circulation. 1998;98:946–952.

8. Praz F, Borger MA, Lanz J, Marin-Cuartas M, Abreu A, Adamo M, Ajmone Marsan N, Barili F, Bonaros N, Cosyns B, et al. 2025 ESC/EACTS Guidelines for the management of valvular heart disease. Eur Heart J. 2025;46:4635–4736.

9. Van Gelder IC, Rienstra M, Bunting KV, Casado-Arroyo R, Caso V, Crijns H, De Potter TJR, Dwight J, Guasti L, Hanke T, et al. 2024 ESC Guidelines for the management of atrial fibrillation developed in collaboration with the European Association for Cardio-Thoracic Surgery (EACTS). Eur Heart J. 2024;45:3314–3414.

10. Rusinaru D, Tribouilloy C, Grigioni F, Avierinos JF, Suri RM, Barbieri A, Szymanski C, Ferlito M, Michelena H, Tafanelli L, et al. Left atrial size is a potent predictor of mortality in mitral regurgitation due to flail leaflets: results from a large international multicenter study. Circ Cardiovasc Imaging. 2011;4:473–481.

11. Sohns C, Marrouche NF. Atrial fibrillation and cardiac fibrosis. Eur Heart J. 2020;41:1123–1131.

12. Grigioni F, Avierinos JF, Ling LH, Scott CG, Bailey KR, Tajik AJ, Frye RL, Enriquez-Sarano M. Atrial fibrillation complicating the course of degenerative mitral regurgitation: determinants and long-term outcome. J Am Coll Cardiol. 2002;40:84–92.

13. Badano LP, Kolias TJ, Muraru D, Abraham TP, Aurigemma G, Edvardsen T, D’Hooge J, Donal E, Fraser AG, Marwick T, et al. Standardization of left atrial, right ventricular, and right atrial deformation imaging using two-dimensional speckle tracking echocardiography: a consensus document of the EACVI/ASE/Industry Task Force to standardize deformation imaging. Eur Heart J Cardiovasc Imaging. 2018;19:591–600.

14. Siebermair J, Kholmovski EG, Marrouche N. Assessment of Left Atrial Fibrosis by Late Gadolinium Enhancement Magnetic Resonance Imaging: Methodology and Clinical Implications. JACC Clin Electrophysiol. 2017;3:791–802.

15. Essayagh B, Antoine C, Benfari G, Messika-Zeitoun D, Michelena H, Le Tourneau T, Mankad S, Tribouilloy CM, Thapa P, Enriquez-Sarano M. Prognostic Implications of Left Atrial Enlargement in Degenerative Mitral Regurgitation. J Am Coll Cardiol. 2019;74:858–870.

16. Dong TX, Li SW, Pan XF, Wang CF, Liu Y, Wu J, Guan XP, Zhang SL, Zuo PF, Liu YL, et al. Normal Values of Echocardiographic Left Atrioventricular Coupling Index and Left Atrial Stiffness Index Reflecting Left Ventricular Diastolic Function: A Multicenter Study. J Am Soc Echocardiogr. 2025;38:794–803.

17. Gal P, Marrouche NF. Magnetic resonance imaging of atrial fibrosis: redefining atrial fibrillation to a syndrome. Eur Heart J. 2017;38:14–19.

18. Suri RM, Clavel MA, Schaff HV, Michelena HI, Huebner M, Nishimura RA, Enriquez-Sarano M. Effect of Recurrent Mitral Regurgitation Following Degenerative Mitral Valve Repair: Long-Term Analysis of Competing Outcomes. J Am Coll Cardiol. 2016;67:488–498.

19. Zoghbi WA, Jone PN, Chamsi-Pasha MA, Chen T, Collins KA, Desai MY, Grayburn P, Groves DW, Hahn RT, Little SH, et al. Guidelines for the Evaluation of Prosthetic Valve Function With Cardiovascular Imaging: A Report From the American Society of Echocardiography Developed in Collaboration With the Society for Cardiovascular Magnetic Resonance and the Society of Cardiovascular Computed Tomography. J Am Soc Echocardiogr. 2024;37:2–63.

20. Sugiura A, Kavsur R, Spieker M, Iliadis C, Goto T, Öztürk C, Weber M, Tabata N, Zimmer S, Sinning JM, et al. Recurrent Mitral Regurgitation After MitraClip: Predictive Factors, Morphology, and Clinical Implication. Circ Cardiovasc Interv. 2022;15:e010895.

21. Thomas JD, Edvardsen T, Abraham T, Appadurai V, Badano L, Banchs J, Cho GY, Cosyns B, Delgado V, Donal E, et al. Clinical Applications of Strain Echocardiography: A Clinical Consensus Statement From the American Society of Echocardiography Developed in Collaboration With the European Association of Cardiovascular Imaging of the European Society of Cardiology. J Am Soc Echocardiogr. 2025;38:985–1020.

22. Benito EM, Carlosena-Remirez A, Guasch E, Prat-González S, Perea RJ, Figueras R, Borràs R, Andreu D, Arbelo E, Tolosana JM, et al. Left atrial fibrosis quantification by late gadolinium-enhanced magnetic resonance: a new method to standardize the thresholds for reproducibility. Europace. 2017;19:1272–1279.

23. Le Tourneau T, Messika-Zeitoun D, Russo A, Detaint D, Topilsky Y, Mahoney DW, Suri R, Enriquez-Sarano M. Impact of left atrial volume on clinical outcome in organic mitral regurgitation. J Am Coll Cardiol. 2010;56:570–578.

24. Thomas L, Abhayaratna WP. Left Atrial Reverse Remodeling: Mechanisms, Evaluation, and Clinical Significance. JACC Cardiovasc Imaging. 2017;10:65–77.

25. Debonnaire P, Leong DP, Witkowski TG, Al Amri I, Joyce E, Katsanos S, Schalij MJ, Bax JJ, Delgado V, Marsan NA. Left atrial function by two-dimensional speckle-tracking echocardiography in patients with severe organic mitral regurgitation: association with guidelines-based surgical indication and postoperative (long-term) survival. J Am Soc Echocardiogr. 2013;26:1053–1062.

26. Maffeis C, Rossi A, Tafciu E, Rizzo S, De Gaspari M, Giambruno V, Di Nicola V, Luciani GB, Basso C, Ribichini FL. Relationship between left atrial fibrosis and function in a selected cohort undergoing surgery for severe primary mitral regurgitation. Int J Cardiol. 2026;460:134614.

27. Shokeir FA, Abdel Aziz AM, Karam R, Tawfik AI, Salem MA, Elboghdady A. Left atrioventricular coupling in isolated pediatric mitral valve prolapse with preserved ejection fraction and moderate regurgitation. Sci Rep. 2026;16:2608.

28. Afana A, Hudelo J, Gonçalves T, Garot J, Soulat G, Fauvel C, Dacher JN, Coisne A, Pontana F, Bohbot Y, et al. Left and right atrioventricular coupling: state-of-the-art review. Eur Heart J Cardiovasc Imaging. 2026;27:504–514.

29. Oakes RS, Badger TJ, Kholmovski EG, Akoum N, Burgon NS, Fish EN, Blauer JJ, Rao SN, DiBella EV, Segerson NM, et al. Detection and quantification of left atrial structural remodeling with delayed-enhancement magnetic resonance imaging in patients with atrial fibrillation. Circulation. 2009;119:1758–1767.

30. Mahnkopf C, Badger TJ, Burgon NS, Daccarett M, Haslam TS, Badger CT, McGann CJ, Akoum N, Kholmovski E, Macleod RS, et al. Evaluation of the left atrial substrate in patients with lone atrial fibrillation using delayed-enhanced MRI: implications for disease progression and response to catheter ablation. Heart Rhythm. 2010;7:1475–1481.

31. Goette A, Kalman JM, Aguinaga L, Akar J, Cabrera JA, Chen SA, Chugh SS, Corradi D, D’Avila A, Dobrev D, et al. EHRA/HRS/APHRS/SOLAECE expert consensus on atrial cardiomyopathies: definition, characterization, and clinical implication. Europace. 2016;18:1455–1490.

32. Meucci MC, Fortuni F, Galloo X, Bootsma M, Crea F, Bax JJ, Marsan NA, Delgado V. Left atrioventricular coupling index in hypertrophic cardiomyopathy and risk of new-onset atrial fibrillation. Int J Cardiol. 2022;363:87–93.

33. Stassen J, van Wijngaarden AL, Butcher SC, Palmen M, Herbots L, Bax JJ, Delgado V, Ajmone Marsan N. Prognostic value of left atrial reservoir function in patients with severe primary mitral regurgitation undergoing mitral valve repair. Eur Heart J Cardiovasc Imaging. 2022;24:142–151.

34. Rubiś PP, Dziewięcka E, González A, Cleland JGF. High variability in assays of blood markers of collagen turnover in cardiovascular disease: Implications for research and clinical practice. Eur J Heart Fail. 2025;27:901–904.

35. Świątko M, Baran JM, Czernicka A, Dudek Ł, Szewczyk M, Pietruszka J, Łazarowicz Ł, Kochman W, Dziedzic EA. Myocardial Fibrosis in Cardiovascular Disease: An Integrative Biomarker-Imaging Framework Linking Molecular Mechanisms to Structural Phenotypes. J Clin Med. 2026;15.

